# Baricitinib in patients admitted to hospital with COVID-19 (RECOVERY): a randomised, controlled, open-label, platform trial and updated meta-analysis

**DOI:** 10.1101/2022.03.02.22271623

**Authors:** RECOVERY Collaborative Group, Peter W Horby, Jonathan R Emberson, Marion Mafham, Mark Campbell, Leon Peto, Guilherme Pessoa-Amorim, Enti Spata, Natalie Staplin, Catherine Lowe, David R Chadwick, Christopher Brightling, Richard Stewart, Paul Collini, Abdul Ashish, Christopher A Green, Benjamin Prudon, Tim Felton, Anthony Kerry, J Kenneth Baillie, Maya H Buch, Jeremy N Day, Saul N Faust, Thomas Jaki, Katie Jeffery, Edmund Juszczak, Marian Knight, Wei Shen Lim, Alan Montgomery, Andrew Mumford, Kathryn Rowan, Guy Thwaites, Richard Haynes, Martin J Landray

## Abstract

**Background:** We evaluated the use of baricitinib, a Janus kinase (JAK) 1/2 inhibitor, for the treatment of patients admitted to hospital because of COVID-19.

**Methods:** This randomised, controlled, open-label platform trial (Randomised Evaluation of COVID-19 Therapy [RECOVERY]), is assessing multiple possible treatments in patients hospitalised for COVID-19. Eligible and consenting patients were randomly allocated (1:1) to either usual standard of care alone (usual care group) or usual care plus baricitinib 4 mg once daily by mouth for 10 days or until discharge if sooner (baricitinib group). The primary outcome was 28-day mortality assessed in the intention-to-treat population. A meta-analysis was conducted that included the results from the RECOVERY trial and all previous randomised controlled trials of baricitinib or other JAK inhibitor in patients hospitalised with COVID-19. The RECOVERY trial is registered with ISRCTN (50189673) and clinicaltrials.gov (NCT04381936).

**Findings:** Between 2 February 2021 and 29 December 2021, 8156 patients were randomly allocated to receive usual care plus baricitinib versus usual care alone. At randomisation, 95% of patients were receiving corticosteroids and 23% receiving tocilizumab (with planned use within the next 24 hours recorded for a further 9%). Overall, 513 (12%) of 4148 patients allocated to baricitinib versus 546 (14%) of 4008 patients allocated to usual care died within 28 days (age-adjusted rate ratio 0·87; 95% CI 0·77-0·98; p=0·026). This 13% proportional reduction in mortality was somewhat smaller than that seen in a meta-analysis of 8 previous trials of a JAK inhibitor (involving 3732 patients and 425 deaths) in which allocation to a JAK inhibitor was associated with a 43% proportional reduction in mortality (rate ratio 0.57; 95% CI 0.45-0.72). Including the results from RECOVERY into an updated meta-analysis of all 9 completed trials (involving 11,888 randomised patients and 1484 deaths) allocation to baricitinib or other JAK inhibitor was associated with a 20% proportional reduction in mortality (rate ratio 0.80; 95% CI 0.71-0.89; p<0.001). In RECOVERY, there was no significant excess in death or infection due to non-COVID-19 causes and no excess of thrombosis, or other safety outcomes.

**Interpretation:** In patients hospitalised for COVID-19, baricitinib significantly reduced the risk of death but the size of benefit was somewhat smaller than that suggested by previous trials. The total randomised evidence to date suggests that JAK inhibitors (chiefly baricitinib) reduce mortality in patients hospitalised for COVID-19 by about one-fifth.

**Funding:** UK Research and Innovation (Medical Research Council) and National Institute of Health Research (Grant ref: MC_PC_19056).

## INTRODUCTION

In patients admitted to hospital with severe COVID-19, the host immune response is thought to play a key role in driving an acute inflammatory process resulting in hypoxic respiratory failure that may require mechanical ventilator support or lead to death.^1,2^ It has previously been shown that the use of dexamethasone and other corticosteroids reduces the risk of death in patients with severe hypoxic COVID-19 and that the addition of an interleukin-6 (IL-6) receptor blocker further reduces the risk of death in these patients.^3–6^

Baricitinib is an inhibitor of Janus kinase (JAK)1 and JAK2 that is licensed in the UK for the treatment of rheumatoid arthritis and atopic dermatitis. The JAKs are a family of four transmembrane protein kinases (JAK1, JAK2, JAK3 and TYK2) that mediate intracellular signalling of a range of extracellular cytokines and interferons.^7^ JAK inhibition prevents downstream phosphorylation and hence activation of signal transducers and activators of transcription (STAT). Since the JAK-STAT pathway mediates the effect of several cytokines, including IL-6, that are raised in severe COVID-19, JAK inhibitors have been proposed as a potential therapeutic option for severe COVID-19.^8,9^ Baricitinib also has moderate inhibitory activity against tyrosine kinase 2 (TYK2) and genetic data support a causal link between high TYK2 expression and life-threatening COVID-19.^10^ Baricitinib was also predicted, using artificial intelligence, to reduce endocytosis of SARs-CoV-2 into lung cells by inhibiting AP2-associated protein kinase 1 (AAK1) and cyclin G associated kinase (GAK).^8^

Baricitinib was tested in combination with remdesivir in the Adaptive Covid-19 Treatment Trial-2 (ACTT-2) and was shown to improve time to recovery compared to remdesivir alone (rate ratio for recovery 1.16, 95% CI 1.01-1.32). There was also a suggestion that 28-day mortality may be reduced by baricitinib (HR 0.65, 95% CI 0.39-1.09).^11^ As a consequence, the US Food and Drug Administration (US FDA) issued an emergency use authorisation for the use of baricitinib in combination with remdesivir, for the treatment of COVID-19 in hospitalised patients requiring oxygen, invasive mechanical ventilation or extracorporeal membrane oxygenation (ECMO).^12^ Since then, a further 7 randomised trials of JAK inhibitors have reported,^13–19^ of which two have reported a significant reduction in mortality.^16,19^ Here we report the results of a large randomised controlled trial of baricitinib in patients hospitalised with severe COVID-19.

## METHODS

### Study design and participants

The Randomised Evaluation of COVID-19 therapy (RECOVERY) trial is an investigator-initiated, individually randomised, controlled, open-label, platform trial to evaluate the effects of potential treatments in patients hospitalised with COVID-19. Details of the trial design and results for other possible treatments (dexamethasone, hydroxychloroquine, lopinavir-ritonavir, azithromycin, tocilizumab, convalescent plasma, colchicine, aspirin, and casirivimab plus imdevimab) have been published previously.^3,5,20–26^ The trial is underway at 177 hospital organisations in the United Kingdom supported by the National Institute for Health Research Clinical Research Network (appendix pp 3-27). Of these, 159 UK hospitals took part in the evaluation of baricitinib. The trial is coordinated by the Nuffield Department of Population Health at the University of Oxford (Oxford, UK), the trial sponsor. The trial is conducted in accordance with the principles of the International Conference on Harmonisation–Good Clinical Practice guidelines and approved by the UK Medicines and Healthcare products Regulatory Agency (MHRA) and the Cambridge East Research Ethics Committee (ref: 20/EE/0101). The protocol and statistical analysis plan are available in the appendix (pp 61-140) with additional information available on the study website www.recoverytrial.net.

Patients aged at least 2 years admitted to hospital were eligible for the study if they had clinically suspected or laboratory confirmed SARS-CoV-2 infection and no medical history that might, in the opinion of the attending clinician, put the patient at significant risk if they were to participate in the trial. Patients were ineligible for the comparison of baricitinib vs. usual care if aged <2 years, eGFR <15 mL/min/1.73m^2^ or on dialysis or haemofiltration, neutrophil count <0.5 × 10^9^/L, had evidence of active TB infection, or were pregnant or breastfeeding. Written informed consent was obtained from all patients, or a legal representative if patients were too unwell or unable to provide consent.

### Randomisation and masking

Baseline data were collected using a web-based case report form that included demographics, level of respiratory support, major comorbidities, suitability of the study treatment for a particular patient, and treatment availability at the study site (appendix pp 36-38). Data on SARS-CoV-2 vaccination status were collected from 22 December 2020.

Eligible and consenting patients were assigned in a 1:1 ratio to either usual standard of care plus baricitinib or usual standard of care alone, using web-based simple (unstratified) randomisation with allocation concealed until after randomisation (appendix pp 29-33). For some patients, baricitinib was unavailable at the hospital at the time of enrolment or was considered by the managing physician to be either definitely indicated or definitely contraindicated. These patients were excluded from the randomised comparison between baricitinib versus usual care. Patients allocated to baricitinib were to receive baricitinib 4 mg daily for 10 days (or until discharge if sooner). The dose was to be reduced for patients with eGFR <60 mL/min/1.73m^2^ or receiving probenecid, and for children aged <9 years (see appendix p 28 for dosing details). Prior or subsequent administration of tocilizumab was permitted at the discretion of the managing doctor who was also responsible for considering the risk of infection and gastrointestinal perforation (particularly in the context of corticosteroid use).

As a platform trial, and in a factorial design, patients could be simultaneously randomised to other treatment groups: i) colchicine versus usual care, ii) aspirin versus usual care, iii) dimethyl fumarate versus usual care, iv) casirivimab+imdevimab versus usual care, and v) empagliflozin versus usual care. Further details of when these factorial randomisations were open are provided in the supplementary appendix (pp 29-33). Participants and local study staff were not masked to the allocated treatment. The Trial Steering Committee, investigators, and all other individuals involved in the trial were masked to outcome data during the trial.

### Procedures

An online follow-up form was completed by site staff when patients were discharged, had died, or at 28 days after randomisation, whichever occurred first (appendix pp 39-46). Information was recorded on adherence to allocated trial treatment, receipt of other COVID-19 treatments, duration of admission, receipt of respiratory or renal support, new cardiac arrhythmia, thrombosis, clinically significant bleeding, non-COVID infection, and vital status (including cause of death). In addition, routinely collected healthcare and registry data were obtained, including information on vital status at day 28 (with date and cause of death); discharge from hospital; and receipt of respiratory support or renal replacement therapy.

### Outcomes

Outcomes were assessed at 28 days after randomisation, with further analyses specified at 6 months. The primary outcome was 28-day all-cause mortality. Secondary outcomes were time to discharge from hospital, and, among patients not on invasive mechanical ventilation at randomisation, the composite outcome of invasive mechanical ventilation (including extra-corporeal membrane oxygenation) or death. Prespecified subsidiary clinical outcomes were use of invasive or non-invasive ventilation among patients not on any ventilation at randomisation, time to successful cessation of invasive mechanical ventilation (defined as cessation of invasive mechanical ventilation within, and survival to, 28 days), and use of renal dialysis or haemofiltration. Prespecified safety outcomes were cause-specific mortality, major cardiac arrhythmia, thrombotic and major bleeding events, and other infections. Information on suspected serious adverse reactions was collected in an expedited fashion to comply with regulatory requirements. Details of the methods used to ascertain and derive outcomes are provided in the appendix (pp.141-161).

### Statistical Analysis

For all outcomes, intention-to-treat analyses compared patients randomised to baricitinib with patients randomised to usual care. Through the play of chance in the unstratified randomisation, patients in the baricitinib group were slightly older than patients in the usual care group (Table 1). In accordance with the prespecified statistical analysis plan for dealing with baseline imbalances in important prognostic factors (appendix p 125), estimates of the effect of allocation to baricitinib on major outcomes were adjusted for age in three groups (<70 years, ≥70 to <80 years, and ≥80 years). (Exploratory analyses were conducted without this adjustment and, separately, with further adjustment for other predefined subgroups of interest.)

**Table 1:**
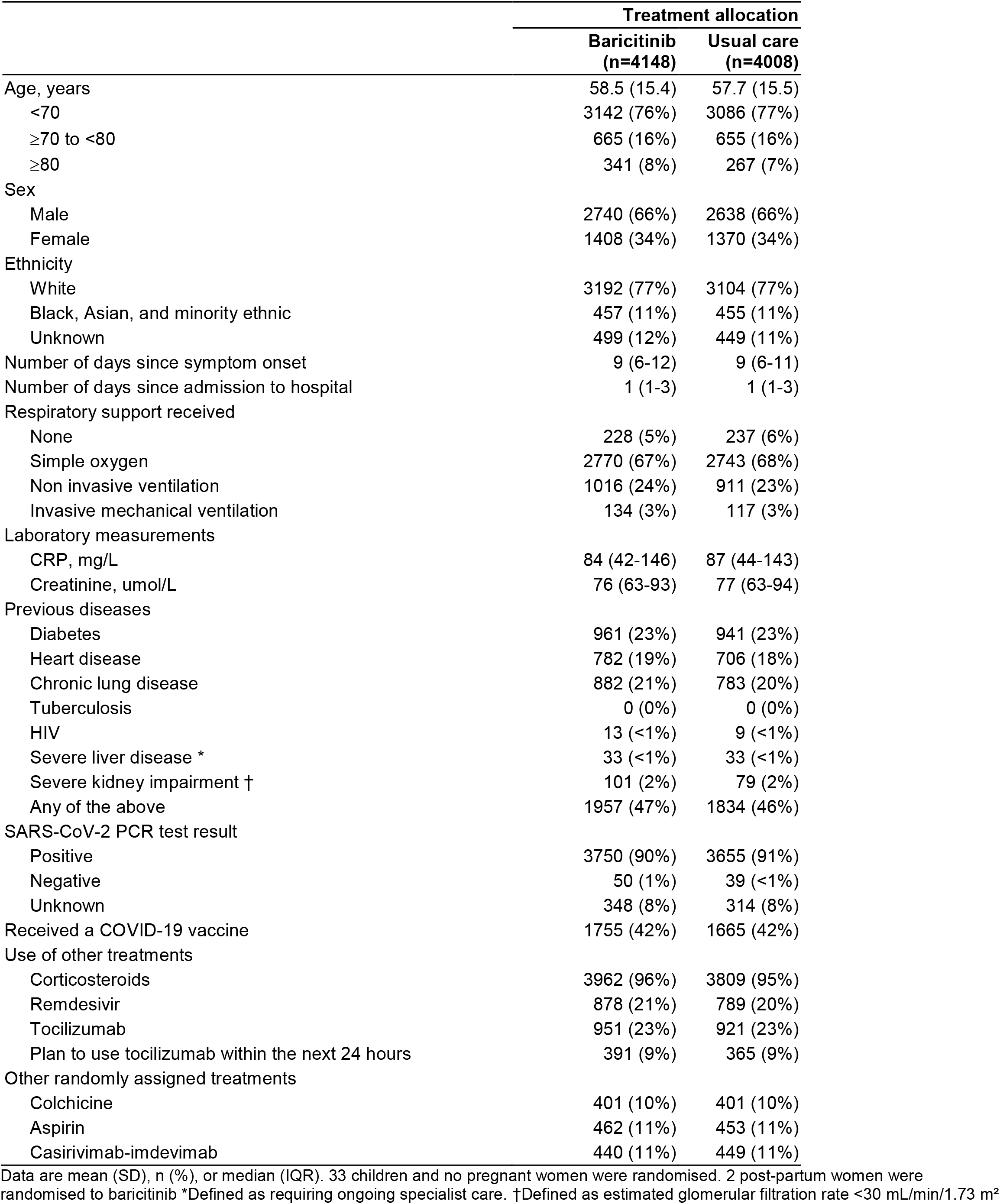
Baseline characteristics by treatment allocation.

For the primary outcome of 28-day mortality, the hazard ratio from an age-adjusted Cox model was used to estimate the mortality rate ratio. We constructed Kaplan-Meier survival curves to display cumulative mortality over the 28-day period. We used the same method to analyse time to hospital discharge and successful cessation of invasive mechanical ventilation, with patients who died in hospital right-censored on day 29. Median time to discharge was derived from Kaplan-Meier estimates. For the pre-specified composite secondary outcome of progression to invasive mechanical ventilation or death within 28 days (among those not receiving invasive mechanical ventilation at randomisation), and the subsidiary clinical outcomes of receipt of ventilation and use of haemodialysis or haemofiltration, the precise dates were not available and so a log-binomial regression model was used to estimate the age-adjusted risk ratio. Estimates of rate and risk ratios (both denoted RR) are shown with 95% confidence intervals.

Prespecified analyses of the primary outcome were done in subgroups defined by six characteristics at the time of randomisation (age, sex, ethnicity, days since symptom onset, level of respiratory support, and use of corticosteroids) with tests of heterogeneity or trend, as appropriate. The full database is held by the study team which collected the data from study sites and performed the analyses at the Nuffield Department of Population Health, University of Oxford (Oxford, UK).

The independent Data Monitoring Committee reviewed unblinded analyses of the study data and any other information considered relevant to the trial at intervals of around 2 to 4 weeks (depending on speed of enrolment) and was charged with determining if, in their view, the randomised comparisons in the study provided evidence on mortality that was strong enough (with a range of uncertainty around the results that was narrow enough) to affect national and global treatment strategies (see appendix p 50).

As stated in the protocol, appropriate sample sizes could not be estimated when the trial was being planned. On the advice of the Trial Steering Committee, recruitment to this comparison was closed on 29 December 2021 when over 8150 patients had been randomised and the blinded 28-day mortality rate was 12.9% (suggesting there would be at least 1050 deaths), giving at least 90% power to detect a proportional risk reduction in the primary outcome of one-fifth at 2P=0.01. The Trial Steering Committee and all other individuals involved in the trial were masked to outcome data until after the close of recruitment.

For the primary outcome of 28-day mortality, the results from the RECOVERY trial were subsequently included in a meta-analysis of results from all previous randomised controlled trials of a JAK inhibitor for patients hospitalised with COVID-19. Details of the systematic search methods are provided in the appendix (p 28). For each trial, we compared the observed number of deaths among patients allocated to the JAK inhibitor with the expected number if all patients were at equal risk (i.e., we calculated the observed minus expected statistic [o–e], and its variance v). For RECOVERY, these were estimated from the age-adjusted mortality log rate ratio and its standard error but for other trials, where the exact timing of each death was not available, these were calculated from standard formulae for 2 × 2 contingency tables. We then combined trial results using the log of the mortality rate ratio calculated as the inverse-variance-weighted average S/V with variance 1/V (and hence with 95% CI S/V ±1·96/√V), where S is the sum over all trials of (o–e) and V is the sum over all trials of v.^27^ Such meta-analyses do not make any assumptions about the nature of any true heterogeneity in the log RR between different trials (in particular it does not assume that it is zero). Analyses were performed using SAS version 9.4 and R version 4.0.3. The trial is registered with ISRCTN (50189673) and clinicaltrials.gov (NCT04381936).

## RESULTS

Between 2 February 2021 and 29 December 2021, 8156 (75%) of 10852 patients enrolled into the RECOVERY trial at one of the 169 participating sites were eligible to be randomly allocated to baricitinib (i.e. the treatment was available in the hospital at the time and the attending clinician was of the opinion that the patient had no known indication for or contraindication to it, figure 1). 4148 patients were randomly allocated to baricitinib and 4008 were randomly allocated to usual care. The mean age of study participants in this comparison was 58.1 years (SD 15.5) with a chance imbalance whereby patients randomly allocated to baricitinib were, on average, 0.8 years older than those allocated usual care group (Table 1). At randomisation, 7771 (95%) patients were receiving corticosteroids, 1872 (23%) were receiving tocilizumab (with planned use within the next 24 hours recorded for a further 756 [9%]) (Table 1, Webtable 1). About two-thirds were receiving simple oxygen and one quarter were receiving non-invasive ventilation, with small numbers receiving invasive mechanical ventilation or no respiratory support at all. 3420 (42%) patients had received at least one dose of a SARS-CoV-2 vaccine.

**Figure 1:**
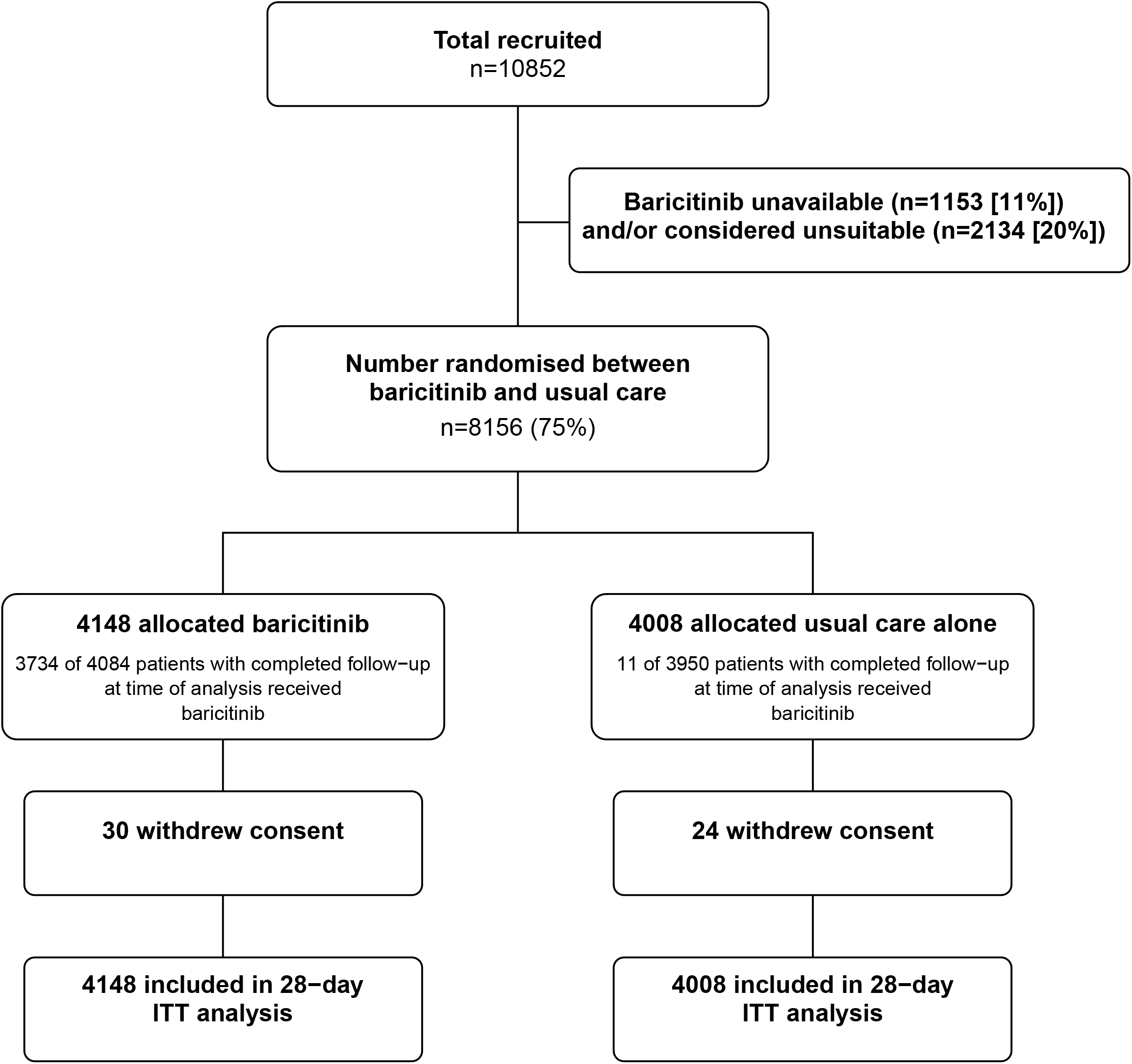
Trial profile. ITT=intention to treat. Baricitinib unavailable and baricitinib unsuitable groups are not mutually exclusive. *Number recruited overall during the period that adult participants could be recruited into the baricitinib comparison.

The follow-up form was completed for 4084 (98%) patients in the baricitinib group and 3950 (99%) patients in the usual care group. Among patients with a completed follow-up form, 91% allocated to baricitinib were reported to have received the treatment compared with <1% allocated to usual care (figure 1, webtable 2). Use of other treatments for COVID-19 was broadly similar among patients allocated baricitinib and among those allocated usual care, with nine-tenths receiving a corticosteroid, one-fifth receiving remdesivir, and one-tenth receiving casirivimab+imdevimab, although use of tocilizumab during the follow-up period was slightly lower in the baricitinib group than in the usual care group (26% vs. 29%) (webtable 2).

Primary and secondary outcome data are known for >99% of randomly assigned patients. Allocation to baricitinib was associated with a significant reduction in the primary outcome of 28-day mortality compared with usual care alone: 513 (12%) of 4148 patients in the baricitinib group died vs 546 (14%) of 4008 patients in the usual care group (age-adjusted rate ratio 0·87; 95% CI 0·77–0·98; p=0·026; table 2, figure 2). Similar proportional risk reductions were seen in sensitivity analyses adjusted for all pre-specified subgroups (as listed in figure 3) and without adjustment for the 0.8 year age-imbalance between randomised groups (webtable 3), and when restricted to participants with a positive SARS-CoV-2 PCR test (age adjusted RR 0.90, 95% CI 0.79-1.02).

**Table 2:**
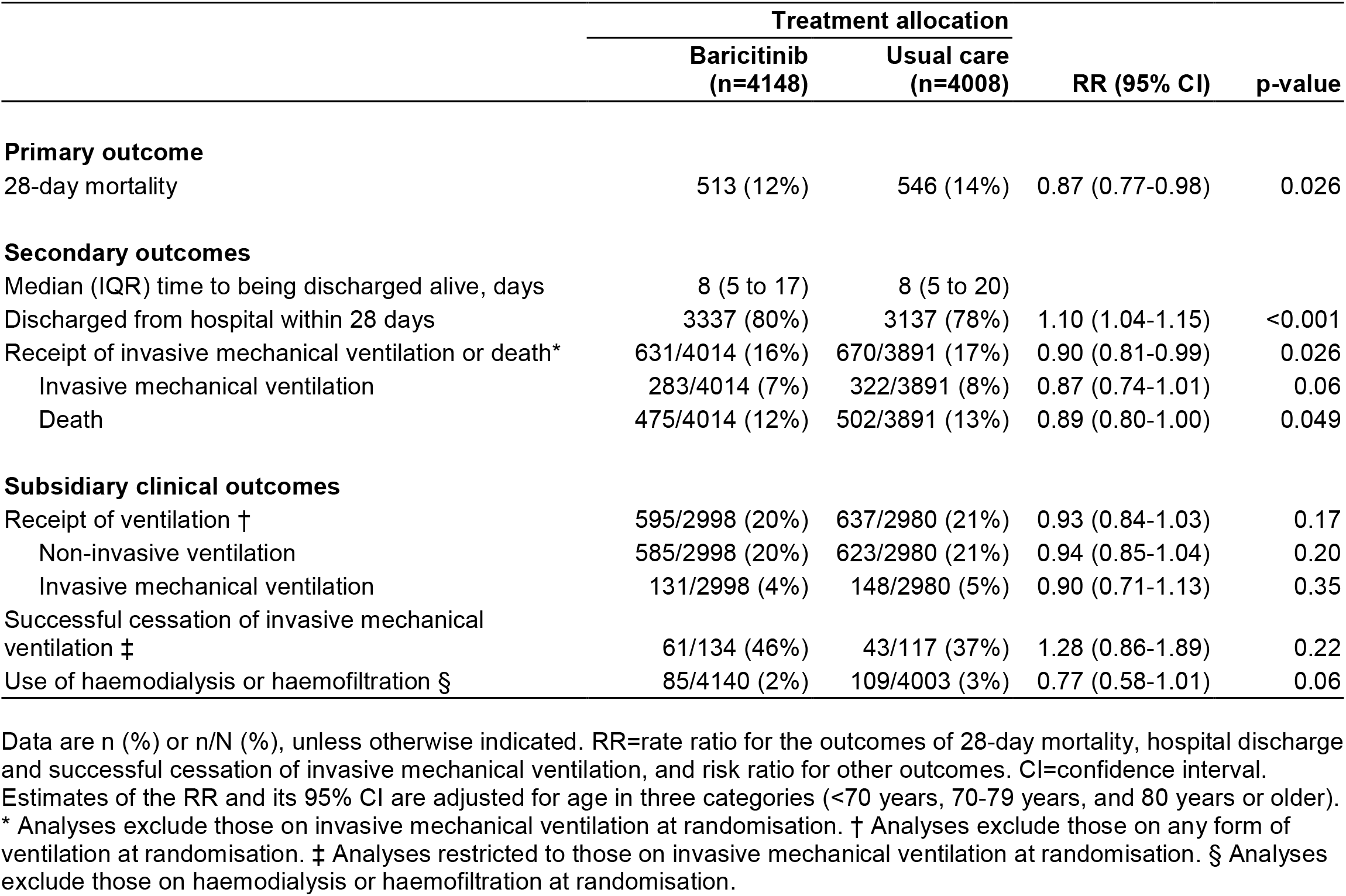
Effect of allocation to baricitinib on key study outcomes.

**Figure 2:**
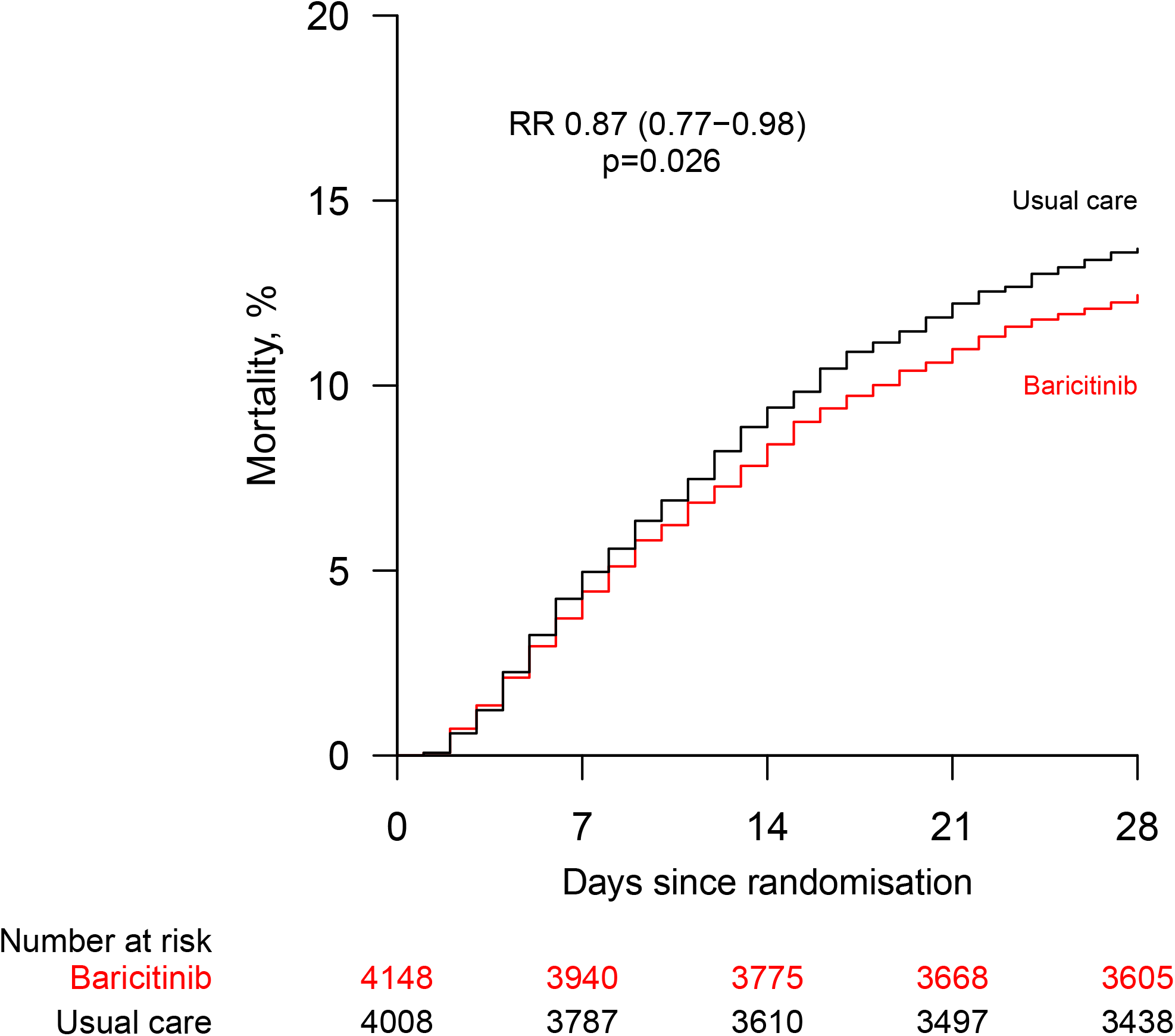
Effect of allocation to baricitinib on 28–day mortality. RR=age-adjusted rate ratio

**Figure 3:**
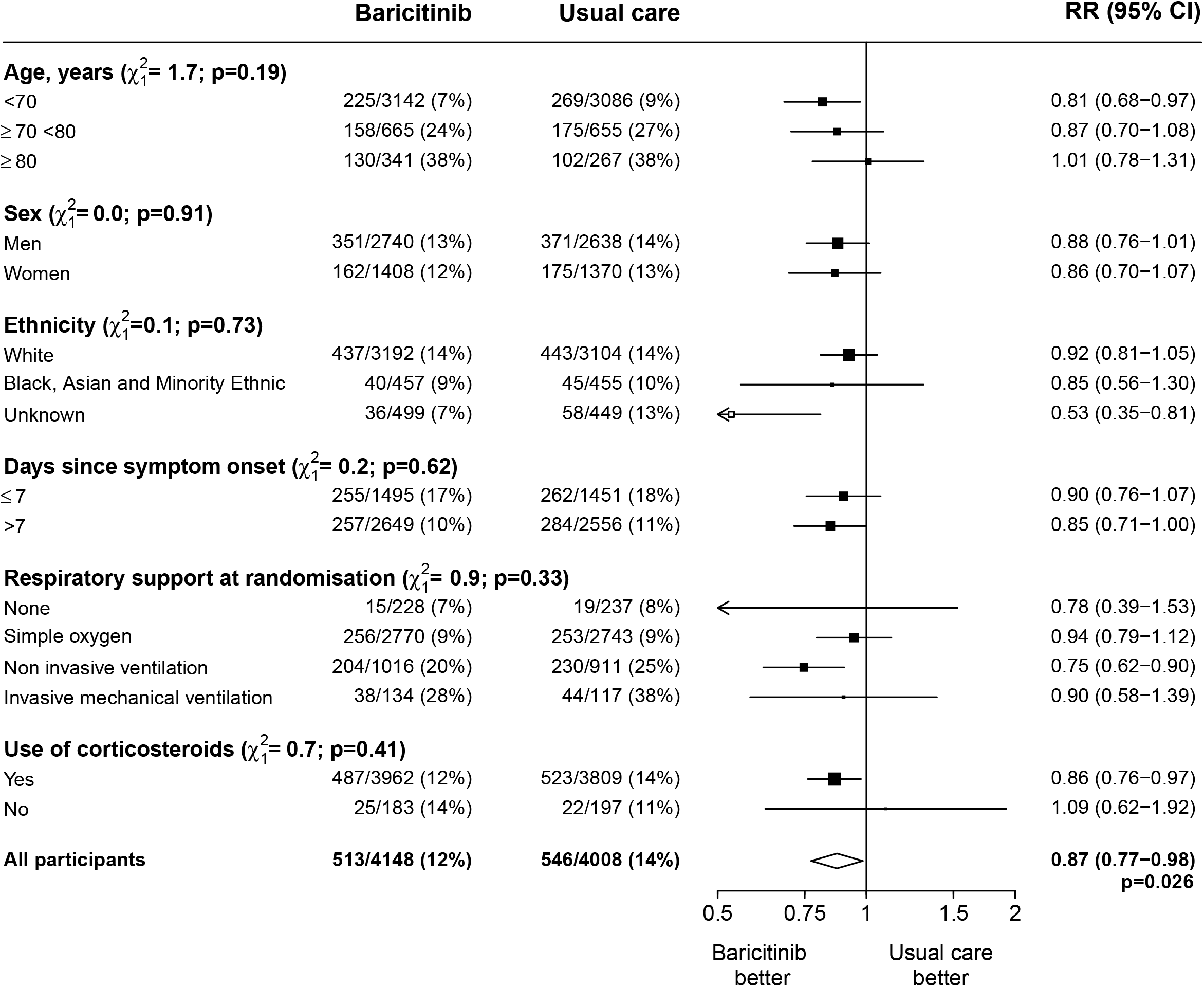
Effect of allocation to baricitinib on 28–day mortality by pre-specified baseline characteristics. Subgroup–specific rate ratio estimates are represented by squares (with areas of the squares proportional to the amount of statistical information) and the lines through them correspond to 95% CIs. The days since onset and use of corticosteroids subgroups exclude patients with missing data, but these patients are included in the overall summary diamond. RR=age adjusted rate ratio.

The proportional effect of baricitinib on mortality was consistent across all pre-specified subgroups (Figure 3), including by level of respiratory support received (test for trend p=0.33), use of dexamethasone at randomisation (test for heterogeneity p=0.41) and, in exploratory analyses, by baseline CRP level (test for trend p=0.93) or by use of tocilizumab or remdesivir at baseline (tests for heterogeneity p=0.53 and p=0.12, respectively) (webfigure 1). There was no evidence that the effect of baricitinib on mortality varied depending on concurrent randomised allocation to colchicine, aspirin or casirivimab+imdevimab (all interaction p-values >0.32).

Discharge alive within 28 days was more common among those allocated to baricitinib compared with usual care (80% vs. 78%; age-adjusted rate ratio 1·10, 95% CI 1·04 to 1·15; median 8 days [IQR 5 to 17] vs. 8 days [IQR 5 to 20]) (table 2 and webfigure 2). Among patients not on invasive mechanical ventilation at baseline, allocation to baricitinib was associated with a lower risk of progressing to the composite secondary outcome of invasive mechanical ventilation or death (16% vs. 17%, age-adjusted risk ratio 0·90, 95% CI 0·81 to 0·99) (table 2 and webfigure 3). The proportional effects of baricitinib versus usual care on these secondary outcomes were also similar across all pre-specified subgroups (webfigures 2 and 3).

There were no significant differences in the pre-specified subsidiary clinical outcomes of cause-specific mortality other than that due to COVID-19 (Webtable 4) or in use of ventilation, successful cessation of invasive mechanical ventilation, or receipt of haemodialysis or haemofiltration (table 2). There were no significant differences in the rates of non-coronavirus infection, thrombotic events, or clinically significant bleeding, but allocation to baricitinib was associated with a nominally significant reduction in new onset cardiac arrythmia (2.3% vs 3.1%, p=0.017) (webtable 5). There were 13 reports of a serious adverse reaction believed to be related to treatment with baricitinib (webtable 6), including 5 participants with a serious non-COVID infection, 3 with a bowel perforation and 2 with a pulmonary embolism.

Our systematic search identified 8 previous trials of a JAK inhibitor for the treatment of patients hospitalised with COVID-19, involving a total of 3732 randomised patients and 425 deaths (figure 4 and webtable 8). ^11,13–19^ In these 8 trials, allocation to a JAK inhibitor was associated with a significant 43% proportional reduction in mortality (rate ratio 0.57; 95% CI 0.45-0.72). This was significantly greater than the mortality risk reduction seen in RECOVERY (test for heterogeneity, p=0.001). After inclusion of the results from RECOVERY into this meta-analysis, the average mortality rate ratio from all 9 trials, now involving 11,888 randomised patients and 1484 deaths, was 0.80 (0.71-0.89; p<0.001).

**Figure 4:**
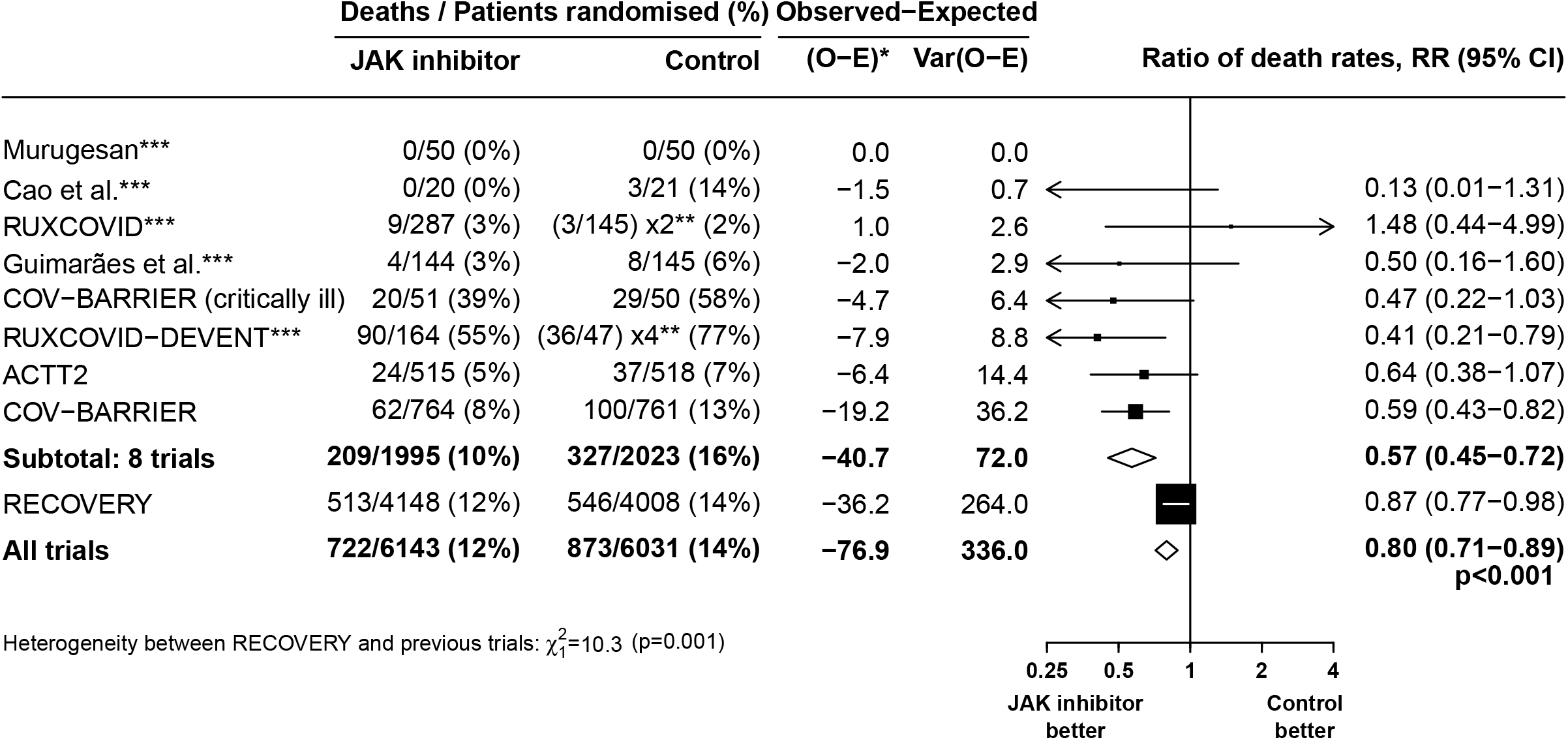
JAK inhibitor vs usual care in patients hospitalised with COVID – Meta-analysis of mortality in RECOVERY and other trials. O-E=observed-expected. Var=variance. RR=Ratio of death rates. Details of the individual studies, including the use of placebo or other treatments in the control group are shown in Webtable 7. * For RECOVERY, the O-E and its variance are calculated from the age-adjusted log RR and its standard error. For the other trials the O-E statistics and their variances are calculated from 2×2 tables. RR is calculated by taking ln RR to be (O-E)/V with Normal variance 1/V. Subtotals or totals of (O-E) and of V yield inverse-variance-weighted averages of the ln RR values without making any assumptions about the true heterogeneity in RRs between trials. ** For balance, controls in the n:1 studies count n times in the control totals and subtotals, but count only once when calculating their O-E and V values. *** These trials did not assess baricitinib but assessed another JAK inhibitor. Restricting the meta-analysis to RECOVERY plus the 3 other baricitinib trials the RR would be 0.81 (95% CI 0.73-0.91).

## DISCUSSION

In this large, randomised trial, allocation to baricitinib significantly reduced 28-day mortality by about one-eighth. This is somewhat less than had been suggested by eight previous randomised, controlled trials of a JAK inhibitor which, together, suggested that allocation to a JAK inhibitor in COVID-19 patients reduces 28-day mortality by about two-fifths. RECOVERY was more than three times the size (in terms of statistical information) of these 8 previous trials put together. When combined in an updated meta-analysis, allocation to baricitinib or another JAK inhibitor in these 9 trials was associated with a significant reduction in 28-day mortality of one-fifth. Although not as large as perhaps previously thought, this still represents an important reduction in mortality risk for patients hospitalised because of COVID-19.

Strengths of the RECOVERY trial included that it was randomised, had a large sample size, broad eligibility criteria and more than 99% of patients were followed up for the primary outcome. The study has some limitations: this randomised trial is open label (i.e., participants and local hospital staff are aware of the assigned treatment), however, the outcomes are unambiguous and were ascertained without bias through linkage to routine health records. Use of tocilizumab during the follow-up period was slightly lower among those allocated to baricitinib compared with control (26% vs. 29%). Based on what we already know about the effects of tocilizumab, would, if anything, lead to a small underestimate of the effects of baricitinib. Furthermore, use of anti-viral or immunomodulatory treatments known to reduce mortality in this setting was similar in those allocated baricitinib and those allocated usual care. Information on radiological, virological or physiological outcomes was not collected.

The smaller effect size observed in RECOVERY compared to earlier trials of baricitinib may simply be a chance effect. However, several other factors could have contributed. The patient population in RECOVERY may have been broader than some of the other trials, which may have been enriched for patients more likely to benefit from immunomodulatory therapy. The use of concomitant therapies has varied between the trials. For example, the ACTT-2 trial did not permit the use of dexamethasone as a treatment for COVID-19, and ACTT-2, COV-BARRIER, RUXCOVID and the study by Guimaraes and colleagues all excluded the use of an IL-6 receptor blocker.^11,15,16,18^ Other factors that may be different between the trials include the prevalence of SARS-CoV-2 vaccination and the predominant circulating SARS-CoV-2 variant. However, there is no reason to believe that, among patients admitted to hospital with severe COVID-19 requiring oxygen or ventilatory support, the proportional risk reduction in mortality with baricitinib, a host-directed therapy, would differ by vaccination status or SARS-CoV-2 variant. Despite the heterogeneity of effect between RECOVERY and the previous 8 trials combined, the overall result of the meta-analysis (which makes no assumptions about the nature of any true differences in treatment effects between the different populations studied) provides the best guide of the proportional benefits that might be expected from the use of baricitinib in clinical practice.

The size of the RECOVERY trial allows exploration of the effects of treatment among different subgroups of patients. The benefits of baricitinib on 28-day mortality were consistent across all subgroups, including by age, sex, ethnicity, C-reactive protein, and level of respiratory support received (although over 90% participants were either on simple oxygen or receiving non-invasive mechanical ventilation). The benefits of baricitinib were also consistent regardless of concomitant treatment with remdesivir, a systemic corticosteroid and/or an IL-6 receptor blocker (tocilizumab or sarilumab). Reassuringly, we found no evidence that allocation to baricitinib was associated with excess rates of non-COVID mortality, non-COVID infection or thrombosis by comparison with usual care.

In November 2020, the US FDA granted Emergency Use Authorisation for baricitinib in combination with remdesivir and it has yet to receive marketing approval for COVID-19.^12^ US National Institutes of Health guidelines updated in February 2022 recommend the use of baricitinb for patients on dexamethasone who have rapidly increasing oxygen needs and systemic inflammation.^28^ In January 2022, the World Health Organization updated their COVID-19 therapeutics guidelines to include a strong recommendation for the use of baricitinib as an alternative to an IL-6 receptor blocker, in combination with corticosteroids, in patients with severe or critical COVID-19.^29^ The results from the RECOVERY trial and our meta-analysis, considerably strengthen the evidence that baricitinib can reduce mortality and other adverse clinical outcomes in patients hospitalised with COVID-19 and support the co-administration of baricitinib with dexamethasone and/or an IL-6 receptor blocker.

In summary, this large, randomised trial confirms evidence from previous smaller trials that treatment with baricitinib can reduce mortality in patients hospitalised with COVID-19, although the size of the benefit is about half that previously thought. The benefits appear to be consistent regardless of treatment with remdesivir, systemic corticosteroids and/or an IL-6 receptor blocker such as tocilizumab. The results support the use of baricitinib in addition to other immunosuppressive therapies in patients hospitalised with COVID-19.

### Research in context

#### Evidence before this study

We searched Medline, Embase, MedRxiv, bioRxiv and the WHO International Clinical Trials Registry Platform from Sept 1, 2019 to Feb 13, 2022, for randomised controlled trials evaluating the effect of bariticinib or another Janus Kinase (JAK) inhibitor in patients hospitalised with COVID-19 using the search terms (“SARS-CoV-2.mp” OR “SARS-CoV2” OR “SARSCoV2.mp” OR “COVID.mp” OR “COVID-19.mp” OR “COVID19.mp” OR “2019-nCoV.mp” OR “Coronavirus.mp” or “Coronavirinae/”) AND (“JAK inhibitor.mp or Janus kinase inhibitor/” OR “Janus kinase inhibitor.mp” OR “Baricitinib.mp or baricitinib/” OR terms for other specific JAK inhibitors (listed in appendix p 28) and using validated filters to select for randomised controlled trials. No language restrictions were applied.

We identified eight relevant randomised trials with results available that assessed JAK inhibitors in hospitalised patients with COVID-19: three assessed baricitinib, three assessed ruxolitinib and two assessed tofacitinib. Six of the trials had been fully published of which four were considered to be low risk of bias for the 28-day mortality outcome with two having some concerns (one because of lack of information about pre-specified analyses and some imbalances between randomised groups of other interventions given during the trial; the other because of lack of information about the randomisation process, inconsistency in reporting of outcome endpoint timing and lack of information about pre-specified analyses). A meta-analysis of these eight trials, which included a total of 425 deaths among 3732 patients, suggested that allocation to a JAK inhibitor was associated with a 43% proportional reduction in 28-day mortality (OR 0.57 [95% confidence interval 0.45-0.72]).

#### Added value of this study

The Randomised Evaluation of COVID-19 Therapy (RECOVERY) trial is the largest randomised trial of the effect of a JAK inhibitor in patients hospitalised with COVID-19. We found that in 8156 patients admitted to hospital with COVID-19, baricitinib reduced 28-day mortality by 13%, increased the probability of discharge alive within 28 days, and, amongst patients who were not receiving invasive mechanical ventilation at randomisation, reduced the probability of progression to the composite outcome of invasive mechanical ventilation or death. The benefits were consistent in all subgroups of patients, including those receiving a systemic corticosteroid and/or an IL-6 receptor blocker.

#### Implications of all the available evidence

The randomised evidence from all 9 completed JAK inhibitor trials to date suggest that treatment with baricitinib or an alternative JAK inhibitor reduces mortality by about one-fifth in hospitalised COVID-19 patients, including those already receiving a systemic corticosteroid and/or an IL-6 receptor blocker.

## Data Availability

The protocol, consent form, statistical analysis plan, definition & derivation of clinical characteristics & outcomes, training materials, regulatory documents, and other relevant study materials are available online at www.recoverytrial.net. As described in the protocol, the trial Steering Committee will facilitate the use of the study data and approval will not be unreasonably withheld. Deidentified participant data will be made available to bona fide researchers registered with an appropriate institution within 3 months of publication. However, the Steering Committee will need to be satisfied that any proposed publication is of high quality, honours the commitments made to the study participants in the consent documentation and ethical approvals, and is compliant with relevant legal and regulatory requirements (e.g. relating to data protection and privacy). The Steering Committee will have the right to review and comment on any draft manuscripts prior to publication. Data will be made available in line with the policy and procedures described at: https://www.ndph.ox.ac.uk/data-access. Those wishing to request access should complete the form at https://www.ndph.ox.ac.uk/files/about/data_access_enquiry_form_13_6_2019.docx and e-mailed to

## Contributors

This manuscript was initially drafted by the PWH and MJL, further developed by the Writing Committee, and approved by all members of the Trial Steering Committee. PWH and MJL vouch for the data and analyses, and for the fidelity of this report to the study protocol and data analysis plan. PWH, MM, JKB, MB, LCC, JD, SNF, TJ, EJ, KJ, WSL, AMo, AMu, KR, RH, and MJL designed the trial and study protocol. MM, LP, MC, G P-A, CL, DRC, CB, RS, PC, AA, CAG, BP, TF, AK, and the Data Linkage team at the RECOVERY Coordinating Centre, and the Health Records and Local Clinical Centre staff listed in the appendix collected the data. NS and JRE did the statistical analysis. All authors contributed to data interpretation and critical review and revision of the manuscript. PWH and MJL had access to the study data and had final responsibility for the decision to submit for publication.

## Writing Committee (on behalf of the RECOVERY Collaborative Group)

Peter W Horby,* Jonathan R Emberson,* Marion Mafham,* Mark Campbell, Leon Peto, Guilherme Pessoa-Amorim, Enti Spata, Natalie Staplin, Catherine Lowe, David R Chadwick, Christopher Brightling, Richard Stewart, Paul Collini, Abdul Ashish, Christopher A Green, Ben Prudon, Timothy Felton, Anthony Kerry, J Kenneth Baillie, Maya H Buch, Saul N Faust, Thomas Jaki, Katie Jeffery, Edmund Juszczak, Wei Shen Lim, Alan Montgomery, Andrew Mumford, Kathryn Rowan, Richard Haynes,^+^ Martin J Landray.^+^

^*^ PWH, JRE and MM made an equal contribution

^+^ RH and MJL made an equal contribution

## Data Monitoring Committee

Peter Sandercock, Janet Darbyshire, David DeMets, Robert Fowler, David Lalloo, Mohammed Munavvar, Adilia Warris, Janet Wittes.

## Data sharing

The protocol, consent form, statistical analysis plan, definition & derivation of clinical characteristics & outcomes, training materials, regulatory documents, and other relevant study materials are available online at www.recoverytrial.net. As described in the protocol, the Trial Steering Committee will facilitate the use of the study data and approval will not be unreasonably withheld. Deidentified participant data will be made available to bona fide researchers registered with an appropriate institution within 3 months of publication. However, the Steering Committee will need to be satisfied that any proposed publication is of high quality, honours the commitments made to the study participants in the consent documentation and ethical approvals, and is compliant with relevant legal and regulatory requirements (e.g. relating to data protection and privacy). The Steering Committee will have the right to review and comment on any draft manuscripts prior to publication. Data will be made available in line with the policy and procedures described at: https://www.ndph.ox.ac.uk/data-access. Those wishing to request access should complete the form at https://www.ndph.ox.ac.uk/files/about/data_access_enquiry_form_13_6_2019.docx and e-mailed to

## Acknowledgements

Above all, we would like to thank the thousands of patients who participated in this trial. We would also like to thank the many doctors, nurses, pharmacists, other allied health professionals, and research administrators at 169 NHS hospital organisations across the whole of the UK, supported by staff at the National Institute of Health Research (NIHR) Clinical Research Network, NHS DigiTrials, Public Health England, Department of Health & Social Care, the Intensive Care National Audit & Research Centre, Public Health Scotland, National Records Service of Scotland, the Secure Anonymised Information Linkage (SAIL) at University of Swansea, and the NHS in England, Scotland, Wales and Northern Ireland.

The RECOVERY trial is supported by grants to the University of Oxford from UK Research and Innovation (UKRI) and NIHR (MC_PC_19056), the Wellcome Trust (Grant Ref: 222406/Z/20/Z) through the COVID-19 Therapeutics Accelerator, and by core funding provided by the NIHR Oxford Biomedical Research Centre, the Wellcome Trust, the Bill and Melinda Gates Foundation, the Foreign, Commonwealth and Development Office, Health Data Research UK, the Medical Research Council Population Health Research Unit, the NIHR Health Protection Unit in Emerging and Zoonotic Infections, and NIHR Clinical Trials Unit Support Funding. TJ is supported by a grant from UK Medical Research Council (MC_UU_00002/14). WSL is supported by core funding provided by NIHR Nottingham Biomedical Research Centre. Tocilizumab was provided free of charge for this trial by Roche Products Limited. Regeneron Pharmaceuticals supported the trial through provision of casirivimab and imdevimab. Baricitinib was provided from routine NHS stock. The views expressed in this publication are those of the authors and not necessarily those of the NHS, the NIHR or the Department of Health and Social Care.

